# Breakthrough SARS-CoV-2 infections after COVID-19 mRNA vaccination in MS patients on disease modifying therapies

**DOI:** 10.1101/2021.12.23.21268177

**Authors:** Maria Pia Sormani, Irene Schiavetti, Matilde Inglese, Luca Carmisciano, Alice Laroni, Caterina Lapucci, Valeria Visconti, Carlo Serrati, Ilaria Gandoglia, Tiziana Tassinari, Germana Perego, Giampaolo Brichetto, Paola Gazzola, Antonio Mannironi, Maria Laura Stromillo, Cinzia Cordioli, Doriana Landi, Marinella Clerico, Elisabetta Signoriello, Jessica Frau, Maria Teresa Ferrò, Alessia Di Sapio, Livia Pasquali, Monica Ulivelli, Fabiana Marinelli, Marcello Manzino, Graziella Callari, Rosa Iodice, Giuseppe Liberatore, Francesca Caleri, Anna Maria Repice, Susanna Cordera, Mario Alberto Battaglia, Marco Salvetti, Diego Franciotta, Antonio Uccelli, the CovaXiMS study group

**Author notes:** Corresponding author: **Maria Pia Sormani**, Department of Health Sciences, Via Pastore 1, 16132, University of Genova, Italy. These authors have contributed equally to the manuscript.

## Abstract

**Background:** Patients with Multiple Sclerosis (pwMS) treated with anti-CD20 or fingolimod showed a reduced humoral response to mRNA-based SARS-CoV-2 vaccines, while the degree of such responses is unimpaired and similar in pwMS treated with other disease modifying therapies (DMTs), or untreated. However, the nature of the SARS-CoV-2 vaccine-induced immune response is based also on cellular immunity and there is emerging evidence that anti-SARS-CoV-2 specific CD4 and CD8 T cell responses can be detected after vaccination also in patients with low antibody levels. In this study we aimed to monitor the risk of breakthrough SARS-CoV-2 infection and to identify correlates of reduced protection in frail vaccinated pwMS on different DMTs.

**Methods:** We designed a long term clinical follow-up of the CovaXiMS (Covid-19 vaccine in Multiple Sclerosis), a prospective multicenter cohort study enrolling pwMS scheduled for SARS-CoV-2 vaccination with mRNA vaccines and tested for SARS-CoV-2 antibodies before and after the second vaccine dose. These patients were followed with periodic phone calls up to a mean time of 6 months, and all the SARS-CoV-2 breakthrough infections were registered. The impact of DMTs on cumulative incidence of breakthrough Covid-19 cases was presented by Kaplan-Meier curves. A multivariable logistic model was run to assess factors associated to a higher risk of breakthrough infections.

**Findings:** 1705 pwMS (81.6% BNT162b2 and 18.4% mRNA-1273) had a full vaccination cycle (2 vaccine doses, 21/28 days apart). Of them, 1509 (88.5%) had blood assessment 4 weeks after the second vaccine dose. During follow-up, 23 breakthrough Covid-19 infections (cumulative incidence: 1.5%, SE=0.3%) were detected after a mean of 108 days after the second dose (range, 18-230). Of them, 9 were on ocrelizumab, one on rituximab, 4 on fingolimod, 6 on dimethyl-fumarate, one on teriflunomide, and 2 were untreated. Just two cases (a woman on ocrelizumab and a man on teriflunomide) required hospitalization. The probability to be infected was associated only with SARS-CoV-2 antibody levels measured after 4 weeks from the second vaccine dose (HR=0.63, p=0.007); an antibody level of 660 U/mL was calculated as the cutoff for higher risk of infection.

**Interpretation:** Our data show that the risk of breakthrough SARS-CoV-2 infections is mainly associated with reduced levels of the virus-specific humoral immune response.

**Funding:** FISM [2021/Special-Multi/001]; the Italian Ministry of Health grant ‘Progetto Z844A 5×1000’. Italian Ministry of Health: Ricerca Corrente to IRCCS Ospedale Policlinico San Martino.

## Introduction

Evidence of the effect of vaccination against SARS-CoV-2, mainly on virus-specific serological responses^1-3^ and, to a minor extent, on antigen-specific T cell response^4,5^ in patients with MS (pwMS) treated with disease-modifying therapies (DMTs) is rapidly accumulating. There is wide consensus that the use of anti-CD20 monoclonal antibodies and fingolimod are associated with the lowest serum SARS-CoV-2 antibody concentrations, following the SARS-CoV-2 vaccines, whereas virus-specific humoral immune responses in pwMS on all the other DMTs, or untreated, were high and did not differ significantly from healthy controls ^1-4^. On the other hand, there is also growing evidence that vaccinated pwMS treated with anti-CD20 generated robust virus specific CD4 and CD8 T cell responses^4-5^, while these are slightly reduced in fingolimod treated patients^5^. Indeed, Covid-19 vaccination is less immunogenic in immunocompromising conditions^6^ and there is evidence that both serum SARS-CoV-2 antibody levels and the degree of vaccine-induced protection from Covid-19 decline with time since vaccination^7,8^. Thus, it is mandatory to monitor the incidence of breakthrough infections in pwMS, to better understand the role of humoral and cellular response to SARS-CoV-2 vaccination in preventing Covid-19 and its consequences. A preliminary follow-up study of 344 fully vaccinated people with multiple sclerosis on disease modifying therapy reported 13 breakthrough infections, 10 of which were in patients under anti-CD20 therapy and the remaining 3 on fingolimod^9^.

We planned a clinical follow-up of the pwMS enrolled in the CovaXiMS (Covid-19 vaccine in Multiple Sclerosis)^1^, a prospective multicenter cohort study enrolling Covid-19 vaccinated pwMS, in whom SARS-CoV-2 antibodies were measured before the first and 4 weeks after the second vaccine dose. Aim of the study was to evaluate the incidence of breakthrough infections in relation to baseline characteristics and vaccination-elicited antibody levels, and to identify possible correlates of a reduced protection against the disease and its severe form.

## Patients and Methods

### Study design and participants

This was a clinical follow-up of an observational multicenter prospective study conducted in 35 Italian MS centers on pwMS undergoing the SARS-CoV-2 vaccination. Adult pwMS, with or without a previous SARS-CoV-2 infection, who were scheduled for SARS-CoV-2 vaccination, were included in the study. mRNA vaccines BNT162b2 (Pfizer Inc, and BioNTech), or mRNA-1273 (Moderna Tx, Inc) as per clinical practice and regional indications were allowed. Patients who agreed to provide a first blood test sample just before the vaccination and a second drawing one month after the last dose were enrolled in the study. Patients were then followed up, with monthly phone calls and visits as per clinical practice, and any new SARS-CoV-2 infection, or the occurrence of Covid-19 infection was recorded in a dedicated Case Report Form (CRF).

The study is done in compliance with the principles of the Declaration of Helsinki. The protocol is approved by the regional (CER Liguria: 5/2021 - DB id 11169-21/01/2021) and the centralized national ethical committee AIFA/Spallanzani (Parere n 351, 2020/21). Written informed consent was obtained from all participants before starting any study procedures.

### Study procedures

Eligible subjects were contacted by their neurologist 3 months after the second vaccine dose against Covid-19 and then contacted every month. Data of patients who had a PCR-confirmed Covid-19 diagnosis were collected in a dedicated CRF including information on symptoms and severity of the disease.

### Primary Outcome: breakthrough infection

The primary objective of this analysis was to quantify the incidence of breakthrough SARS-CoV-2 infection among the vaccinated pwMS included in the CovaXiMS study. These conditions entail a PCR-confirmed swab, and a time lag of at least 14 days from the second vaccination dose.

### SARS-CoV-2 antibody measurement

High-affinity pan-Ig antibodies to SARS-CoV-2 were measured by a centralized laboratory with a double-antigen sandwich-based electrochemiluminescence immunoassay (Elecsys®, Roche Diagnostics Ltd, Switzerland). Receptor-binding domain (RBD) antibodies were quantitatively measured to evaluate the humoral immune response to the two RBD-coding mRNA vaccines. RBD antibodies have been shown to positively correlate with SARS-CoV-2 neutralizing antibodies on neutralization assays^10-11^. Serum samples were shipped in dried ice by the centers and stored at -20°C until analysis.

### Statistical analysis

The cumulative incidence of breakthrough infections in the different DMT groups was reported by Kaplan-Meier survival curves. A multivariable Cox model was used to evaluate the impact of DMT class and antibody levels developed 4 weeks after the second vaccination dose on the risk of a breakthrough infection after adjusting for age, sex, BMI, EDSS level, disease duration, presence of comorbidities and vaccine type. The antibody levels were transformed on a Log10 scale, to normalize their distribution and according to previous literature^1^. A ROC curve (run on the subgroup of patients with at least 6 months of follow-up) was used to assess the best antibody level cut-off indicating a protective level against breakthrough infections.

## Results

Data were collected between March 4, 2021 and December 15, 2021. At the time of analysis, we had data on 1705 pwMS (81.6% BNT162b2 and 18.4% mRNA-1273) who had a full vaccination cycle (2 vaccine doses, 21/28 days apart). Of them, 1509 (88.5%) had blood assessment 4 weeks after the second vaccine dose. The median follow-up after the second vaccine dose was 222 days (range, 44-328). The patients’ characteristics and the number of vaccinated patients in each DMT group are reported in Table 1.

**Table 1:** Baseline characteristics of patients with multiple sclerosis who received two vaccine doses.

| Characteristics | Overall (N = 1705) |
| --- | --- |
| **Age, years** (mean, SD) | 46.1 (12.45) |
| **Females** (n, %) | 1161 (68.1%) |
| **BMI, kg/m<sup>2</sup>** (mean, SD) | 24.1 (3.65) |
| **MS phenotype** (n, %) |  |
| *Relapsing remitting* | 1420 (83.3%) |
| *Secondary progressive* | 158 (9.3%) |
| *Primary progressive* | 127 (7.4%) |
| **MS disease duration, years** (median, IQR) | 10.0 [5.0 – 15.6] |
| **EDSS** (median, IQR) | 2.0 [1.0 – 4.0] |
| **MS treatment** (n, %) |  |
| *Ocrelizumab* | 272 (16.0%) |
| *Dimethyl fumarate* | 267 (15.7%) |
| *Natalizumab* | 199 (11.7%) |
| *Interferon* | 192 (11.3%) |
| *Fingolimod* | 173 (10.1%) |
| *Teriflunomide* | 102 (6.0%) |
| *Glatiramer acetate* | 102 (6.0%) |
| *Cladribine* | 51 (3.0%) |
| *Rituximab* | 48 (2.8%) |
| *Alemtuzumab* | 23 (1.3%) |
| *Other* | 26 (1.5%) |
| *Untreated* | 250 (14.7%) |
| **Vaccine product** (n, %) |  |
| *mRNA BNT162b2* | 1347 (79.0%) |
| *mRNA-273* | 314 (18.4%) |
| *Not specified* | 44 (2.6%) |

*MS=Multiple Sclerosis, BMI=Body Mass Index, SD=Standard Deviation, IQR=Interquartile Range, EDSS=Expanded Disability Status Scale.*

**Table 2.**
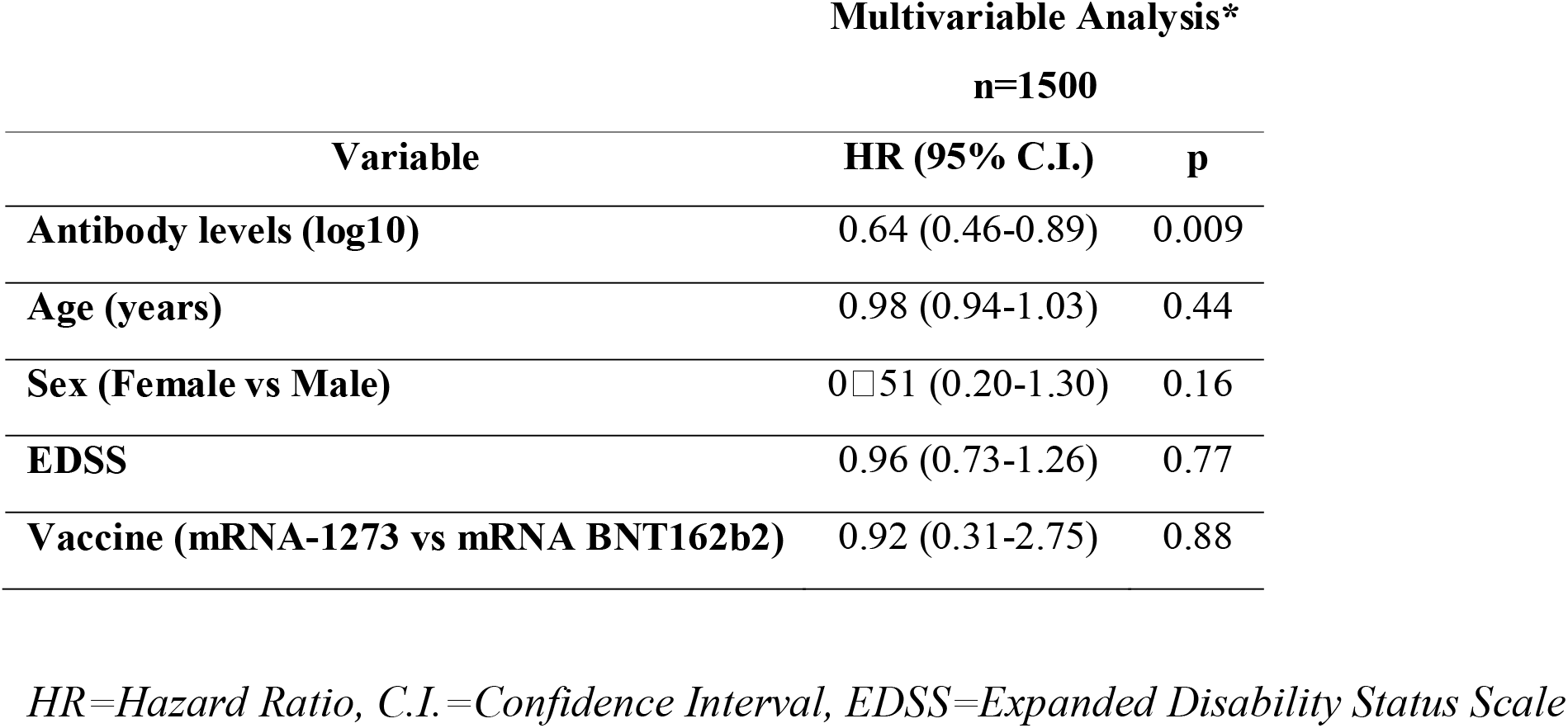
Multivariable Cox regression model evaluating risk factors for breakthrough SARS-CoV-2 infections.

Overall, we observed 23 Covid-19 cases during follow-up reported after a mean of 105 days after the second dose (range, 18-232). Nine of them were on ocrelizumab, one on rituximab, 4 on fingolimod, 6 on dimethyl-fumarate, one on teriflunomide, and 2 were untreated. The 8-month cumulative incidence of breakthrough infection was 1.5% (SE, 0.3%) (Figure 1, panel a). The incidence was higher in pwMS on ocrelizumab (4.0%, SE=1.0%), fingolimod (2.3%, SE=1.2%), rituximab (2.1%, SE=2.2%) or dimethyl-fumarate (2.3%, SE=0.9%), as compared to teriflunomide (1.0%, SE=1.0%), and untreated patients (0.8%, SE, 0.6%), and it was zero for all the other therapies (figure 1, panel b). The log-rank test for heterogeneity among DMTs was borderline significant (p=0.052). Grouping anti-CD20 (ocrelizumab and rituximab) vs fingolimod vs other drugs (Figure 1, panel c) gave a highly significant heterogeneity p<0.001 (Figure 1, panel b).

**Figure 1:**
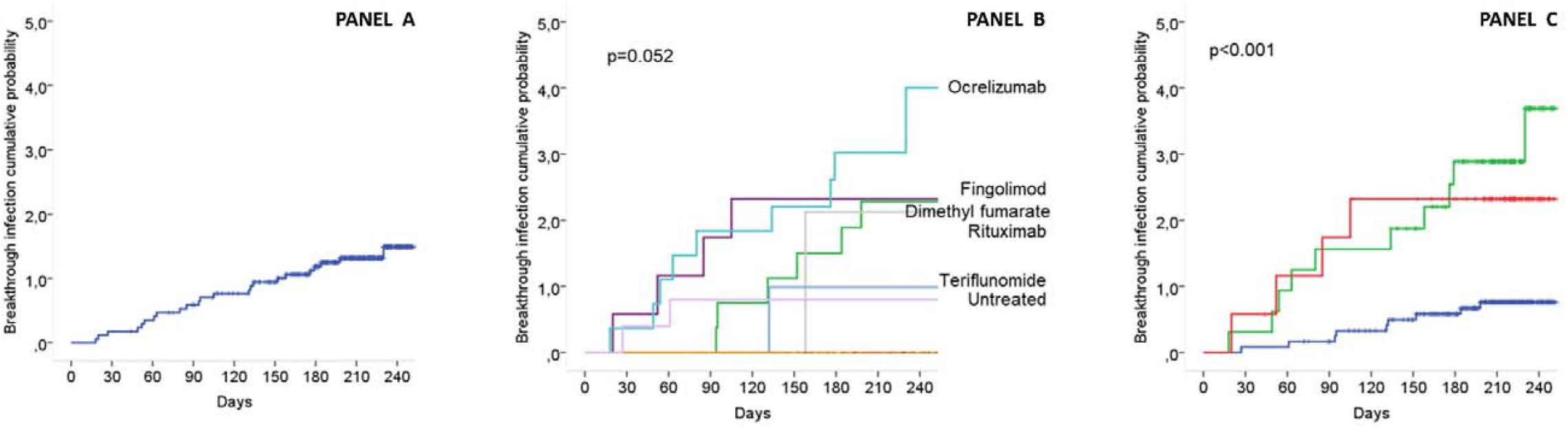
Kaplan Meier survival curves displaying the breakthrough infections cumulative probability overall in the vaccinated cohort (panel a), in groups according to disease modifying therapy (panel b) and in groups according to treatment with anti-CD20, fingolimod and other drugs (panel c).

At multivariable analysis the only significant factor associated to the risk of breakthrough infection was the antibody level after the second dose, with an HR of 0.63 (95%CI=0.45-0.87), p=0.007. This value indicates that the risk of breakthrough infection is reduced by 37% every X10 increase in the antibody level. The ROC curve applied to patients with at least 6 months of follow-up (n=1474, 97.7%) inicated a value of log antibody level of 2.82 (660 U/mL) as the best cutoff discriminating those at a higher risk of infection (AUC=0.71; sensitivity, 88%, specificity, 58%). Figure 2 reports the breakthrough infections in each DMT group and according to antibody levels. Two patients (one in ocrelizumab and one in fingolimod) who had a breakthrough infection, had their blood sample taken after Covid-19. Both patients developed very high antibody levels after Covid-19.

**Figure 2:**
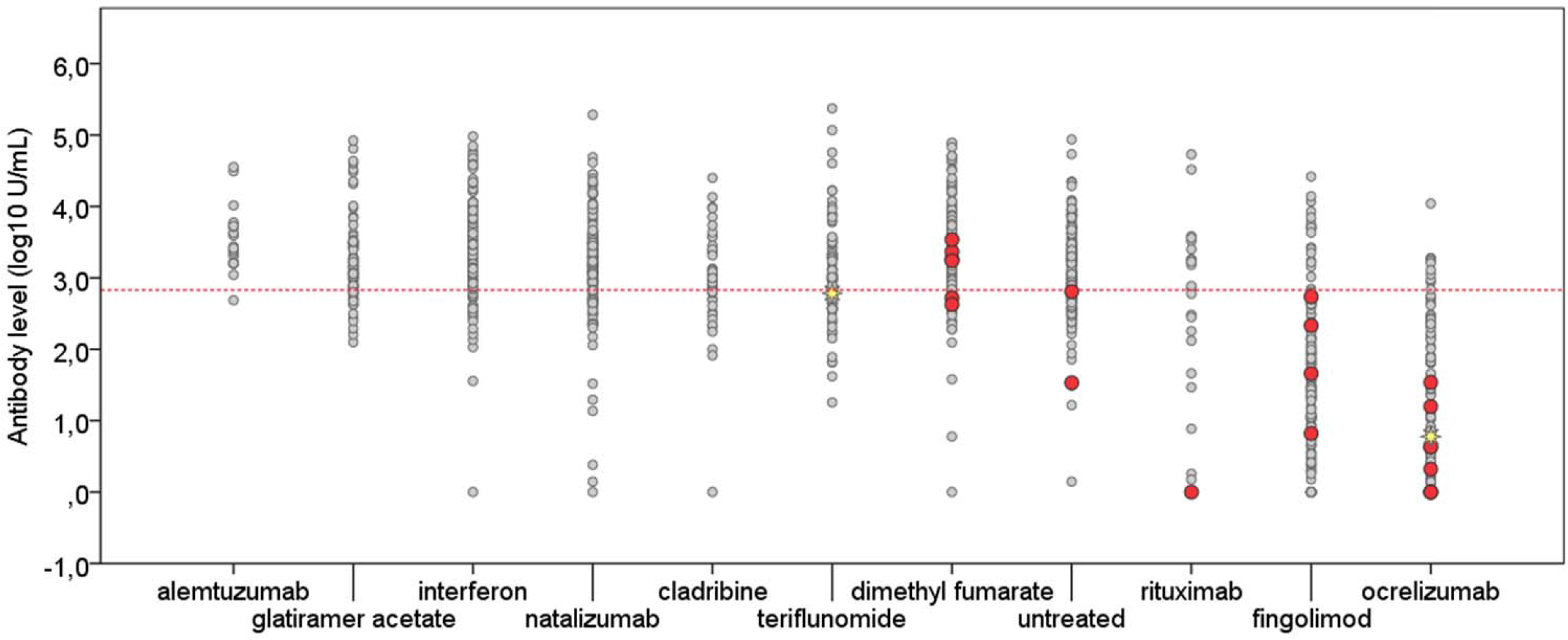
Covid-19 cases (red dots) and hospitalized Covid-19 cases (yellow stars) according to disease modifying therapy and antibody level

Two patients, a woman on ocrelizumab with a very low antibody titer (5 U/mL) and a man on teriflunomide with medium antibody titer (610 U/mL), were hospitalized after Covid-19, but recovered without the need for supplemental oxygen.

## Discussion

The risk of both contracting SARS-CoV-2 infection and of not responding to Covid-19 vaccines is higher in pwMS on anti-CD20 monoclonal antibodies or fingolimod. Vaccine-induced protection from the disease is expected to waning with time since vaccination^7,8^ and different levels of immunity impact on susceptibility to breakthrough infections. In this study we assessed the incidence of breakthrough infections in a large Italian cohort of patients fully vaccinated with mRNA vaccines. The cumulative incidence of breakthrough infection over a follow-up of 8 months was 1.5%, with some heterogeneities among groups treated with different DMTs. However, since the cases are few, a test for heterogeneity among more than 10 groups taking different DMTs has low statistical power. A Cox model, including the antibody level as a continuous variable and adjusted for the main baseline covariates, revealed that lower SARS-CoV-2 antibody levels as the only relevant risk factor for breakthrough infections. The risk of infection decreases of about 37% for every 10 times-fold increase in the antibody levels. In this study, SARS-CoV-2 antibody level equal to, or lower than 660 U/mL are associated with higher risk of infection in the subsequent 6 months. Post-vaccination neutralizing antibody titers predicted the risk of breakthrough infection in health care workers^12^. However, no neutralizing or binding antibody threshold titer identified so far can predict the degree of protection, depending on unpredictable titer changes over time and on the strength of immunity at the moment when a subject is exposed. In addition, other SARS-CoV-2 infection risk modifiers, such as each patient’s safety precautions and viral load exposure amount, can impact on the risk of Covid-19 independently from antibody titers.

On the other hand, the main goal of vaccination is not to prevent infections, but rather to prevent the severe disease. In this respect, we observed just two breakthrough infections that caused Covid-19 requiring hospitalization followed by resolution within one week. The prevalence of hospitalization in our Italian Covid-19 cohort of pwMS in the pre-vaccination era was 12.8%^13^ and therefore around 2 cases were expected out of the 22 infected patients. Therefore, these numbers are too low to draw conclusions from this observation.

Analogously to the other study on anti-CD20 treated patients following Covid-19 vaccination^14^, the age factor, which typically associate with lower antibody responses to Covid-19 vaccines^15^, did not have an independent role on influencing the risk of breakthrough infections in our pwMS cohort, when we take into account the antibody levels. This phenomenon appears to be evident in the general population too^16^.

This study has some limitations. First, incidence data of breakthrough infections in the general population, as well as those of non-breakthrough infections in unvaccinated people, were not available for comparing these rates with those of our pwMS cohort. In solid organ transplant recipients, the diminished antibody responses to SARS-CoV-2 resulted in 41-to-82-fold (depending on the statistical approach) higher risk of breakthrough infection vs general population^17^. Second, we have no data on SARS-CoV-2 molecular characterization, as the Delta variant reduces vaccine effectiveness^17^, but high enough SARS-CoV-2 antibody titers can protect against infection and severe outcomes^18^.

In conclusion, the results of this study suggest that the rates of breakthrough infections in pwMS on DMTs at least partially depends on the level of humoral immunity to SARS-CoV-2, which, is reduced in patients under specific DMTs and that declines over time. Larger cohorts are needed to understand the protective role of vaccination for severe Covid-19 in pwMS under different DMTs. It is likely that the protection from Covid-19 might increase after breakthrough infections, or after subsequent vaccine doses. Identifying the frequency, severity, and predisposing factors of breakthrough infections in frail patients may inform how to protect them with anticipates boosting doses of vaccines. The recommended safety precautions, such as masks and distancing, should remain a mainstay to reduce the incidence of SARS-CoV-2 infection.

## Data Availability

All data produced in the present study are available upon reasonable request to the authors

## Acknowledgements

Supported by FISM [2021/Special-Multi/001]; the Italian Ministry of Health grant ‘Progetto Z844A 5×1000’ to the IRCCS Ospedale Policlinico San Martino (D.F.) and the Italian Ministry of Health: Ricerca Corrente to IRCCS Ospedale Policlinico San Martino. We thank Barbara Uggeri, Claudio Spallarossa, and Giovanni Rossi for their excellent laboratory assistance.

